# Artificial Intelligence Devices for Image Analysis in Digital Pathology

**DOI:** 10.64898/2026.03.23.26349089

**Authors:** GA Matthews, L Godson, C McGenity, D Bansal, D Treanor

## Abstract

**Background:** There is increasing momentum behind the clinical implementation of AI-based software for image analysis in digital pathology. As regulations, standards, and national approaches to the clinical use of AI continue to develop, the marketplace of AI products is expanding and evolving – presenting pathologists with a multitude of devices that offer the potential to improve pathology services.

**Methods:** To maintain pace with this changing AI device landscape, we conducted a comprehensive search for, and analysis of, commercial AI products for image analysis in digital pathology. This included CE-marked and Research Use Only (RUO) products using images with histological stains (e.g., H&E) or immunohistochemical (IHC) labelling. Product information and published clinical validation studies were assessed, to understand the quality of supporting evidence on available products, and product details were compiled into a public register: https://osf.io/gb84r/overview.

**Results:** In total, we identified and assessed 90 CE-marked and 227 RUO AI products. We found that AI products for cancer detection in prostate and breast pathology comprised a substantial portion of the marketplace for H&E image analysis, while IHC products were almost exclusively for use in breast cancer. Clinical validation studies on these products have steadily increased; however, we found that published studies were only available for just over half of H&E products and just over a quarter of IHC products. For CE-marked products, the dataset quality and diversity for AI model performance validation was highly variable, and particularly limited for IHC products. Furthermore, only a limited number of products included studies that assessed measures of clinical utility.

**Conclusion:** As clinical deployment of AI products for image analysis in histopathology grows, there is a need for transparency, rigorous validation, and clear evidence supporting clinical utility and cost-effectiveness. Independent scrutiny of the expanding offering of AI products provides insight into the opportunities and shortcomings in this domain.

## Introduction

The growing implementation of digital pathology in clinical laboratories has spurred interest in image analysis applications that can leverage this digital infrastructure to enhance pathology services. Technological advancements in deep learning (DL)-based image analysis techniques have enabled the development of AI models that demonstrate impressive academic performance in generating diagnostically relevant predictions from digital images. This has led to a flurry of commercialisation of AI-driven products performing diagnostic-support tasks, such as identifying tumour regions, performing tumour grading, quantifying biomarker expression, and estimating patient outcomes^1,2^. Furthermore, national strategies have begun to reflect the mounting optimism that the implementation of AI products will offer a solution to the declining pathology workforce and increasing departmental workload, by improving diagnostic accuracy, workflow efficiency, and patient treatment selection^3,4^.

Despite this promise of clinical benefit, there has so far been a cautious approach to AI deployment^5^. This is partly because pathology departments need to be digitised in order to implement these tools, and although clinical adoption of digital pathology continues to grow, digitised laboratories remain in the minority. There are also regulatory hurdles for AI models of this kind, given their status as medical devices, which has resulted in variable product availability for clinical use across different countries/jurisdictions. In the USA, for example, the FDA has only approved a very small number of devices for histopathology, and limited their use to certain circumstances, with full pathologist oversight with regards to final diagnosis^6,7^. Furthermore, there are significant technical, governance, and economic challenges for clinical institutions to establish the resources, processes, and expertise required to safely deploy and monitor AI technology^8,9^. An additional hesitancy around the deployment of AI tools also stems from the somewhat sparse and variable public evidence supporting the performance and clinical utility of these products in real-world scenarios^10–12^. Progress towards AI deployment, therefore, requires a confidence among clinicians, pathologists, commissioners, and patients that a product’s clinical benefit will outweigh the potential risks, and justify the cost and resources invested in implementation.

While the clinical implementation of AI devices in digital pathology remains in its infancy, there has been continued growth and development within the AI marketplace. As regulations and standards around healthcare AI evolve, and face the challenge of maintaining pace with rapid technological developments (e.g., ^13–15)^, there is a need to independently monitor the offering of AI-based devices and understand the extent of evidence supporting their clinical use. The lack of suitable regulatory databases for AI devices is a known shortcoming across multiple healthcare domains^16^. Certain clinical specialties have tackled this by creating domain-specific registers of products, to track available devices alongside their capabilities and extent of clinical use (e.g.,^17–21)^. In light of the increasing range of products available in pathology, we and others have established independent registers of pathology AI devices for image analysis^10,22^, and included details of their key features and published clinical evidence.

Given the pace of developments in this fast-moving field, and to counter the static nature of many independent registers, we have substantially expanded our original register of products. Here we present a broad analysis of the marketplace of AI devices for image analysis, incorporating a detailed assessment of CE-marked and Research Use Only (RUO) products available for histological stains and immunohistochemical labelling. We examine the key features of these products as well as the evidence available on their clinical validation, the quality and methodology of validation studies, and reported measures of clinical utility. Our findings provide a comprehensive assessment of this evolving field, including insights into the nature of pipeline products, gaps in evidence generation, and opportunities for product alignment with clinical demand.

## Methods

### Identification of AI-based products for digital pathology image analysis

To identify potential vendors of AI-based digital pathology products, we performed a broad grey literature search that incorporated a range of relevant sources. Searches were conducted between 16/5/25 and 4/6/25 with sources including: exhibitors, presenters, and sponsors at major conferences, grant funding awardees, medical device database listings, contributors to pathology Grand Challenges, and advertisements or articles within pathology newsletters, magazines etc. This resulted in an initial list of 564 vendors associated with digital pathology. Vendor websites were then reviewed (between 11/6/25 and 10/9/25) to determine whether they developed AI-based image analysis products. This reduced the list to 75 vendors producing software for AI-based image analysis in digital pathology. The product(s) offered by each vendor were noted and categorised as CE-marked if there was evidence of regulatory approval (e.g., from the vendor website or press releases) and otherwise categorised as Research Use Only (RUO). Products were included if they were described as applying a deep learning (DL) or AI-based image analysis technique to digital pathology images (including Hematoxylin & Eosin (H&E), special stains, immunohistochemistry (IHC), and cytology). Products using Hematoxylin-Eosin-Saffron (HES) images were included within H&E products.

For each product, relevant information was compiled from the company website and other sources if required (e.g. press releases, or third-party platforms such as the Sectra Amplifier Marketplace). The product information compiled included: pathology subspecialty, primary function, model input data required, and EU/USA regulatory approval status. For pathology subspecialty, products were assigned to a ‘Multiple’ category if they could be applied to more than one subspecialty. For primary function, where a product had more than one function described, the most prominent or first listed function was selected. Medical device databases (EUDAMED^23^ and MHRA PARD^24^) were also searched for vendors in August/September 2025, in order to determine vendor/product listings, and other vendor-related information was obtained from their respective LinkedIn pages.

### Data extraction from publications on clinical validation

For CE-market products for histopathology, we identified publications describing their clinical validation through vendor website and by searching publication databases (PubMed, bioRxiv, arXiv, medRxiv, and Google Scholar; searched between 8/9/25 and 15/9/25), using a combination of the product name and company name as search terms. For this analysis, we limited our inclusion to only products using H&E or IHC whole slide images (WSIs) as input, as these represented the largest categories of CE-marked products. Peer-reviewed publications (and preprints where noted) were included if they were available in English and described some aspect of clinical validation of the AI model and/or its clinical utility in a pathology workflow. Conference proceedings, abstracts, or posters were excluded, as were publications describing validation of the AI model for a purpose outside of its intended use (e.g., testing its performance on a type of tissue specimen not covered by existing regulatory approval).

Various information from within each identified publication was extracted and compiled. In instances where a publication described the validation of more than one product, the publication was treated as two separate ‘studies’, and thus there are more studies included in subsequent analyses than unique publications. Studies were categorised as describing internal or external validation, where internal validation was defined as studies where the data used to evaluate model performance originated from the same institution(s) as the training data, and external validation was defined as studies where the data originated from a different institution(s). For the purpose of this analysis, studies where the test dataset was a mixture of data from an external institute(s) and an institute(s) that contributed data to model training/development were categorised as external validation. The independence of studies was also determined. Studies where a vendor employee was an author on the publication, or where the vendor funded the study, were typically classed as not independent. Exceptions were made if a vendor was an author but played no role in the study design or analysis, in which case the study was classed as independent. In addition, exceptions were made if a study was funded or supported by the vendor (through the provision of equipment/software) but the vendor played no role in the study design, data collection, analysis, preparation of the manuscript or decision to publish. Instances where an author received consulting fees for the vendor but had no other vendor-related conflicts were classed as independent.

Details of the validation studies described within publications were extracted by one reviewer and verified by a second reviewer. The extracted details included: the size and composition of validation datasets, reported AI model and reader performance, methodological details relating to ground truth, and reported measures of clinical utility/workflow impact. For each external validation study, details relating to the dataset(s) composition were extracted and aggregated by product. The size of each dataset was defined by the total number of patients/cases and/or WSIs reported. Where only the number of patients/cases was available within a publication, one WSI per case has been assumed for the purpose of analysis. The number of distinct sources of data included within a dataset was defined as the number of separate institutions that samples were obtained from (i.e., if samples originated from two locations, but glass slides were subsequently prepared at one location, this was considered as two data sources). If samples were reported as being obtained from one location but subsequently processed at two different locations, this was considered two distinct sources for the purpose of analysis (in order to take a liberal approach to the breadth of variation in data origin). In addition, we extracted dataset information relating to the number of different countries data was sourced from and the number of scanner models used to generate digital images. Only WSI scanners were included, whereas systems using microscope-mounted cameras were excluded. If the scanner model was not reported in full within a publication, the scanner model was inferred, if possible, from the information provided. If the exact model could not be determined, the scanner was excluded from the analysis of model type, but included in the overall count of distinct scanners used. To aggregate data for each product, where multiple publications were available for a given product, dataset composition values (i.e., size, number of sources, number of countries of origin, and number of scanner models used) were summed to give an overall sense of the size and diversity of data the product had been tested on. If publication details indicated that the data in different studies had originated from the same source or was generated on the same scanner model, these were included only once in the overall total.

In addition to dataset composition, we extracted the reported metrics for standalone AI performance on external datasets, along with associated reader performance where available. The method used to determine the ground truth for external datasets was also extracted and placed into categories. In some cases, multiple datasets were used in a single study, each of which could have different approaches to ground truth determination, so multiple methods from some studies were included. Assigned categories were defined by the number of readers used to determine a consensus diagnosis for each case, as well as whether additional reviewers and/or IHC images were consulted where required. If studies noted that the original diagnosis was used to determine ground truth, it was assumed that additional reviewers (i.e., second opinions) and IHC images would have been available to the reporting pathologist.

## Results

### Regulatory status of AI devices for image analysis

AI devices for image analysis in digital pathology were located by reviewing the websites of 564 companies operating in the digital pathology field. Our broad search and inclusion criteria resulted in the identification of 317 devices from 75 companies, which comprised 90 CE-marked products and 227 RUO products (Figure 1A, Table S1). This included devices using different types of digital image data as input (e.g., H&E, IHC, special stains, cytology images, or a combination of these) as well as devices that required additional clinicopathological information alongside image data to generate a prediction. Additionally, six AI products for image quality control (QC) were identified, however, these are deemed by vendors to fall outside the scope of medical device regulations, and so were not included in either the RUO or CE-marked categories.

**Figure 1.**
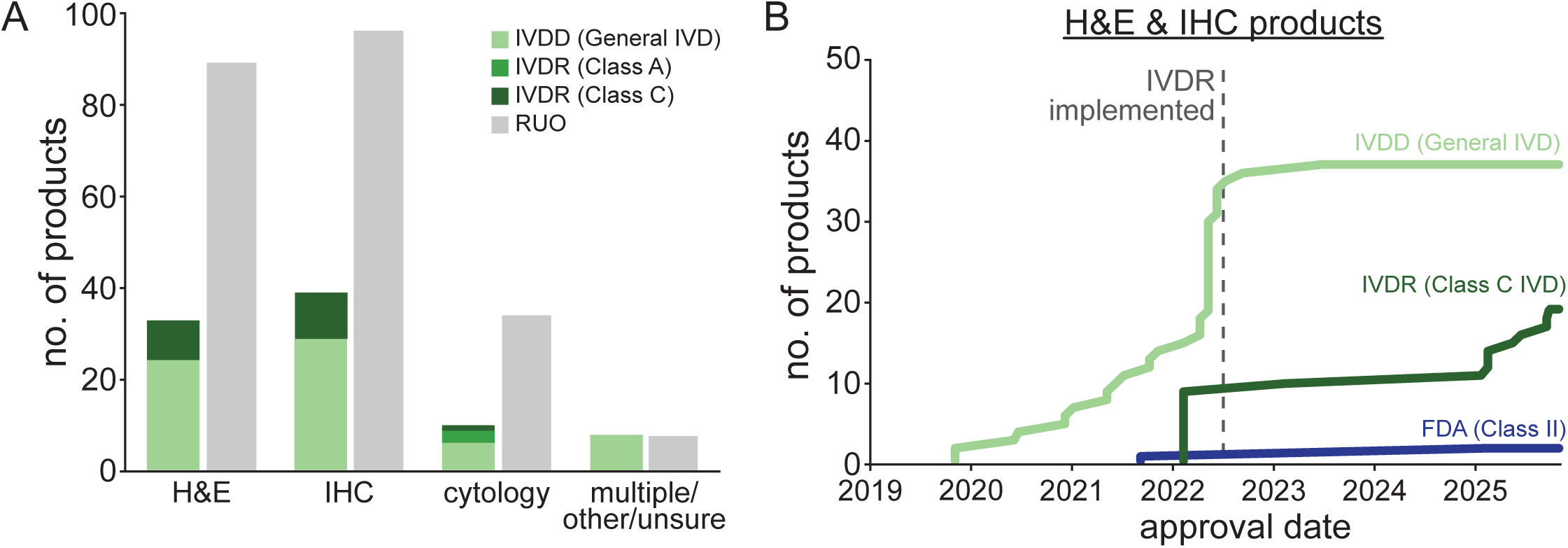
Regulatory status of AI devices for digital pathology image analysis. (A) Bar chart showing the number of devices using H&E, IHC, and cytology images as input, alongside their classification as CE-marked or RUO products. (B) Line graphs indicating the approximate date and type of regulatory approval received for the EU market (i.e., IVDD or IVDR) and the USA market (i.e., FDA) for H&E and IHC products.

Across the 90 devices that were reported to be CE-marked, 33 used H&E images as input, 39 used IHC images, and 10 used cytology images. Of these, 67/90 (74%) had achieved CE-marking under the old IVDD (IVD Directive 98/79/EC) as self-certified General IVDs, while 23/90 (26%) had been approved under the new IVDR (In Vitro Diagnostic Medical Devices Regulation (2017/746)) as Class A or Class C devices (Figure 1A). Where possible, we determined approximate dates of regulatory approval from vendor websites or press releases for H&E and IHC products, which showed that although there was a rapid increase in IVD self-certifications by vendors under IVDD immediately prior to the implementation of the IVDR in May 2022, approvals under the IVDR have begun to slowly, but steadily, increase (Figure 1B). Contrastingly, very few H&E or IHC products have been approved for use by the FDA in the USA (Figure 1B). Considering H&E products, since our prior analysis in 2023^10^, the proportion of devices approved under IVDR has increased from ∼8% to 27%.

We next looked in more depth at the vendors producing AI devices for histopathology image analysis, which showed that over half (62%) of the 29 vendors manufacturing CE-marked devices produced 1-2 devices, 21% 3-4 devices, and 17% >6 products, with a similar distribution for RUO products (Figure S1A). Vendors also tended to focus on solely products for H&E, IHC, or cytology images, with a smaller fraction producing devices for more than one of these image types (21% and 25% of companies for CE-marked and RUO products, respectively; Figure S1B). Vendor size (based on employee number) varied substantially: from SMEs consisting of 2-10 employees to large multinationals with >1,000 employees. Overall, of the 29 vendors manufacturing CE-marked devices, 52% had ≤50 employees, and produced 43% of the identified CE-marked products (Figure S1C-D). Similarly, of the 61 vendors manufacturing RUO devices, 61% had ≤50 employees, and produced 42% of the identified products (Figure S1C-D). Finally, we considered whether devices could be located on medical device databases in the EU and UK. For CE-marked devices, we were able to locate nearly all companies on EUDAMED, but only 20% of companies had their associated device listed (Figure S2A). For placement on the UK market, companies and their devices need to be registered with the MHRA, and we found that about half (51%) of companies with CE-marked devices were listed on the MHRA PARD (Figure S2B).

### Clinical subspecialty and primary function of AI devices

We next assessed the clinical subspecialty and primary function of CE-marked and RUO devices, based on vendor descriptions. For these analyses we focused on AI models using H&E, special stains, or IHC histopathology images as input, and excluded cytology products. For CE-marked devices, the vast majority were intended for use in breast pathology (55%), with a smaller proportion for uropathology (16%), gastrointestinal (12.5%) or cardiothoracic (7.5%) pathology (Figure 2A). When categorised by the type of image input (i.e., H&E or IHC), we found that IHC devices were almost exclusively intended for breast pathology (82%) and cardiothoracic pathology (15%), whereas products for H&E images were dominated equally by products for breast (36%) and uropathology (36%; Figure 2A). There was a similar distribution of subspecialty across RUO products, with 31% of all products and 40% of IHC products for use in breast pathology. However, in contrast to CE-marked devices, there was a greater proportion of products intended for use in other clinical specialties (Figure 2B).

**Figure 2.**
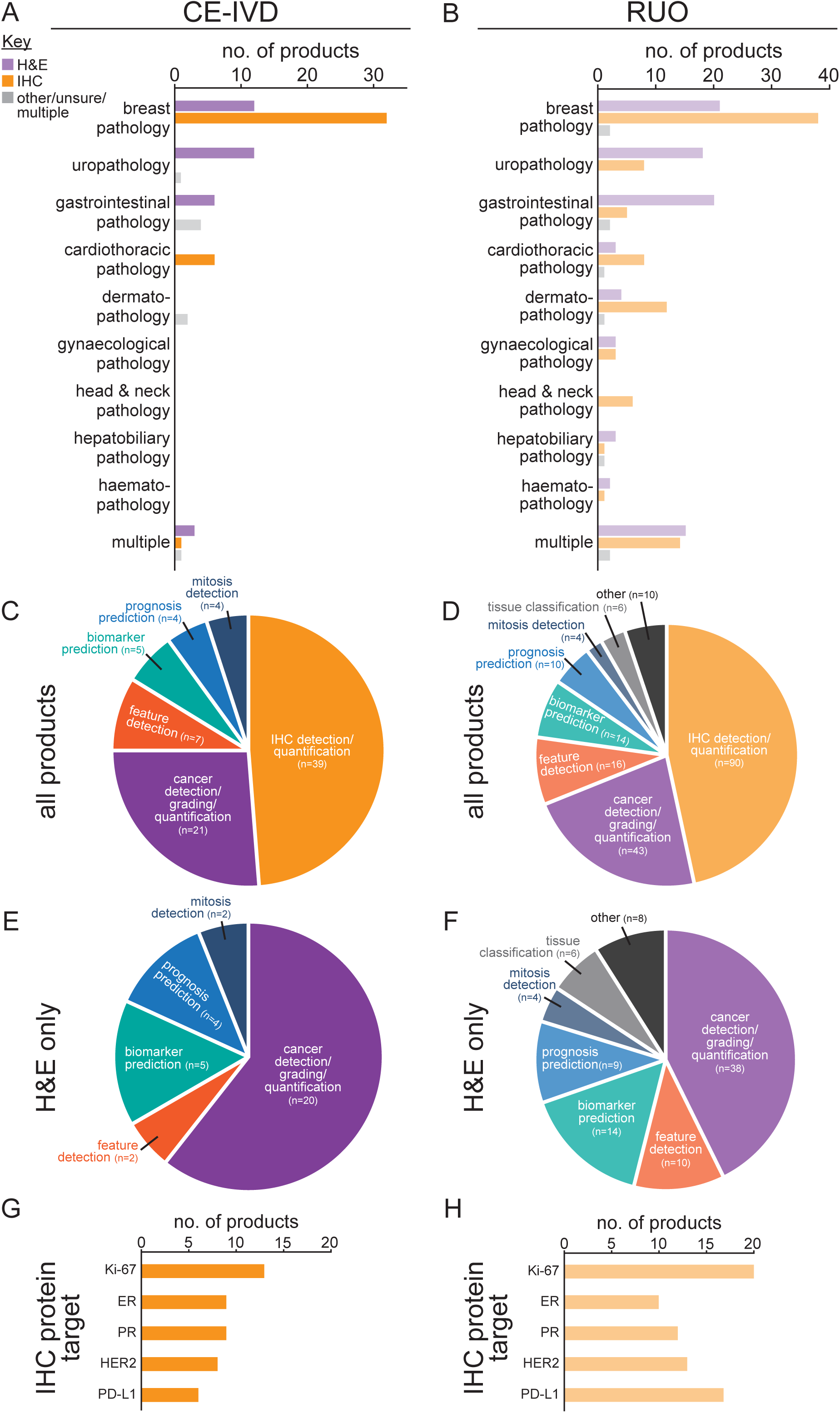
Clinical subspecialty and primary function of AI devices for digital pathology image analysis. (A) Bar charts showing pathology subspecialty of all CE-marked and (B) RUO devices separated by image input type (H&E, IHC, or other). (C) Pie charts showing primary function of all CE-marked and (D) RUO products, and (E-F) only products for H&E image analysis. (G) Top five IHC protein targets analysed by CE-marked and (H) RUO products.

To assess the main purpose of AI devices, their primary function was assigned to one of eight categories. This showed that devices for IHC detection/quantification accounted for approximately half of CE-marked (49%) and RUO (47%) products (Figure 2C-D). When examining only products for H&E image analysis, we found that devices for cancer detection, grading, and/or quantification tasks comprised 61% of all CE-marked H&E products, with a smaller proportion of products focused on biomarker prediction (15%), prognosis prediction (12%), feature detection (6%), and mitosis detection (6%) (Figure 2E). For RUO H&E devices, the proportion of cancer detection models was smaller overall (43%), relative to CE-marked devices, with products focused on other primary functions accounting for a greater proportion of devices (Figure 2F). To further examine devices for IHC image analysis, we looked at the IHC protein target that devices were intended to analyse. The top five protein targets analysed by CE-marked and RUO products were identical, consisting of four breast cancer markers (Ki-67, ER, PR and HER2), and one marker typically used in non-small cell lung cancer (PD-L1) (Figure 2G-H).

Given that breast pathology, uropathology, and gastrointestinal pathology were the most heavily represented subspecialties across CE-marked AI devices, we next compared the features of devices within these subspecialties (Figure S3), alongside approximate case statistics based on data from the UK^25,26^. This showed that while AI models using H&E images were most likely to perform cancer detection across all three subspecialties, there was a wider range of products available for breast pathology and gastrointestinal pathology, compared with uropathology which was more limited to cancer detection (Figure S3A). Furthermore, IHC products were only available for biomarkers used in breast pathology, and not for uropathology nor gastrointestinal pathology. To provide some indication of how this aligns with the impact of cancer cases in these subspecialties, we also plotted the annual case numbers and years of life lost per death, based on UK data, which showed that annual cancer cases for breast, prostate, and gastrointestinal cancers are relatively similar, whereas years lost per death is greatest for breast cancer (Figure S3B).

### Clinical validation of CE-marked AI devices

Clinical validation of AI models is an essential step in determining their performance on unseen data and gives an indication of potential performance in real-world scenarios. However, real-world data is diverse from many respects, and robust validation relies on test datasets being representative of the population and reflecting this diversity. In order to appreciate the quality of clinical validation performed on CE-marked products, we identified publications relating to each product and extracted data on test dataset composition, methodology, and reported results.

For this analysis only CE-marked products using H&E or IHC images were included. We found that publications were available for 18/33 (55%) of H&E products, and 11/39 (28%) of IHC products (Figure 3A-B). In several cases, publications reported the validation of multiple products. These were treated as separate ‘studies’, resulting in a total of 44 studies on H&E products, and 16 studies on IHC products, the majority of which included external validation of model performance. We also assessed the independence of these studies, based on the contribution of the product vendor (see *Methods*), which showed that 10/44 (23%) of studies on H&E products were independent, compared with 9/16 (56%) of studies on IHC products (Figure 3A-B). Overall, of the 18 H&E products with publications, 12 (66%) were associated with 1-2 publications, and six products were associated with ≥3 publications, while for the 11 IHC products with publications, 10 (91%) were associated with 1-2 publications and one product with ≥3 publications (Figure 3C-D).

**Figure 3.**
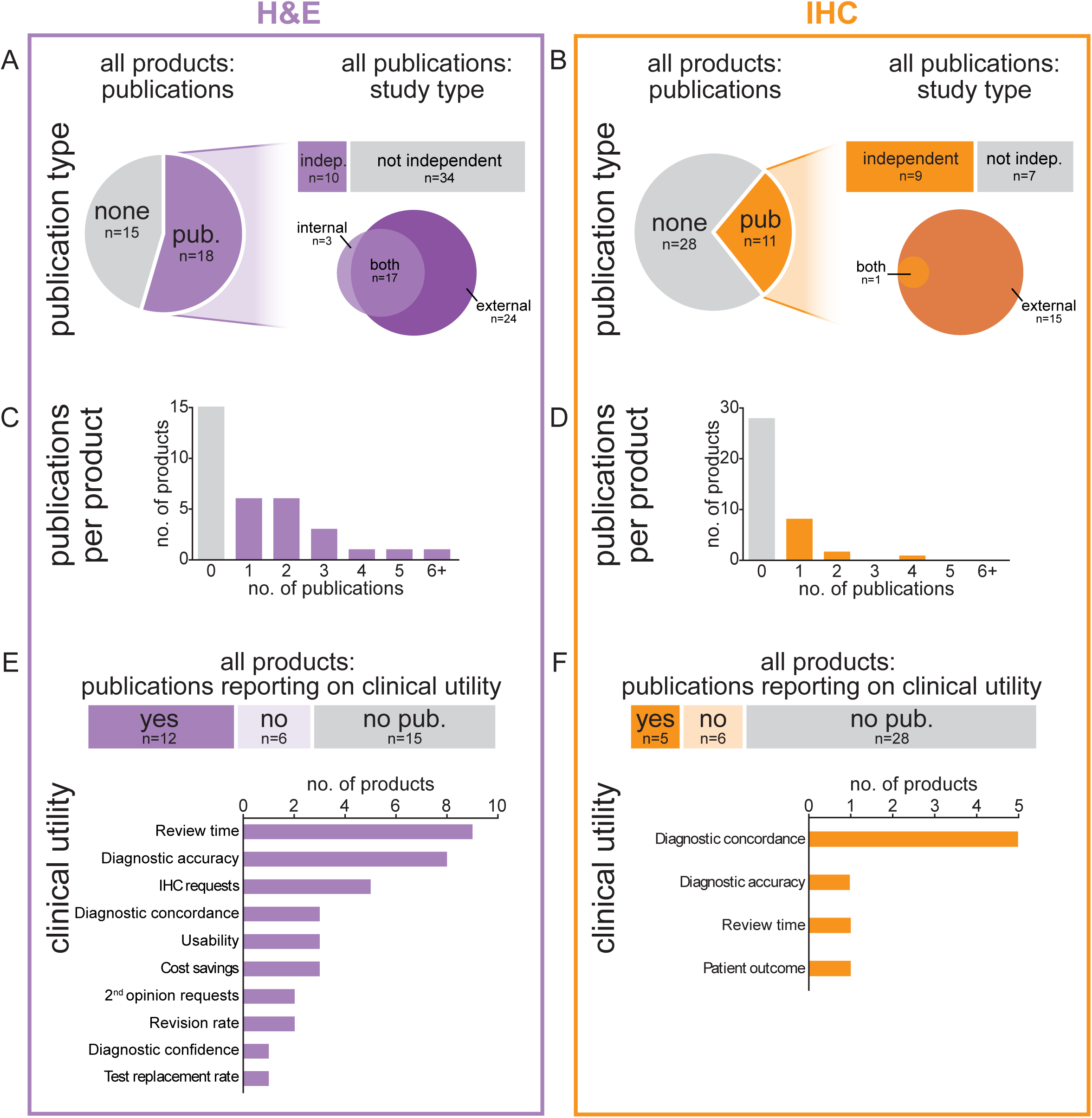
Clinical validation of CE-marked AI devices for digital pathology image analysis. (A) Pie chart showing the proportion of H&E and (B) IHC products with a publication describing AI model clinical validation, and graphs illustrating the vendor independence of those studies and whether they described internal or external validation. Note that, for H&E products, this includes 4 preprints. (C) Bar charts showing the number of publications per product, for models using H&E and (D) IHC images as input. (E) Graphs (upper panels) indicating the proportion of publications that reported on at least one element of clinical utility/workflow impact (beyond standalone AI model accuracy), for H&E and (F) IHC products. Bar graphs (lower panels) show the number of products which report on each workflow category.

Some studies described not only the standalone performance of the AI model on test datasets, but also reported on other measures of clinical utility/workflow impact of using the product. We found that, for H&E products, there were 12/33 products for which at least one publication included an assessment of clinical utility/workflow impact (Figure 3E). The most prevalent categories reported were case review time, diagnostic accuracy, and the number of IHC requests. For IHC products, only 5/39 products had at least one publication reporting on the clinical utility/workflow impact, with diagnostic concordance among pathologists reported most frequently (Figure 3F).

### Composition of data used for external validation of CE-marked AI devices.

External validation is typically used to evaluate AI model performance on data originating from a distinct institution(s) to that used for model training/development. In histopathology, this is particularly important, as patient data obtained from different sources can differ significantly (due to e.g., population demographics, clinical feature presentation, and specimen processing protocols). This can result in variability in the appearance of digital images from different sites, which is known to impact AI model performance. To appreciate the quality of external validation conducted on CE-marked products, we assessed features of the datasets used for model evaluation. As there were instances where multiple external validation studies on each product had been published, we aggregated all studies for each product to provide an indication of the overall extent of validation reported for each product.

When combining data from all external validation studies available on each product, we found that AI models for H&E image analysis had overall reported tested on ∼50-7,000 patients/cases, which equated to ∼50-16,000 WSIs, with the majority of products (12/18) reporting testing on <2,500 WSIs (Figure 4A). In terms of the number of distinct sources of data origin, of the 17 products with published external validation studies, six products reported external validation on 1-3 data sources overall, five products on 4-5 sources, and six products on 8+ sources of data (Figure 4C). The overall extent of reported external validation on AI products for IHC images was significantly lower, with reporting product testing on ∼50-500 patients/cases, equating to ∼50-600 WSIs (Figure 4B-B). For the 11 products with published external validation studies, half (6/11) of these products reported external validation on just one data source overall, four products on 2-5 sources, and one product on seven sources of data (Figure 4D). In terms of the country of data origin, sources based in the USA featured most frequently in the external validation datasets for H&E and IHC products, followed by countries based in Europe (Figure 4E-F).

**Figure 4.**
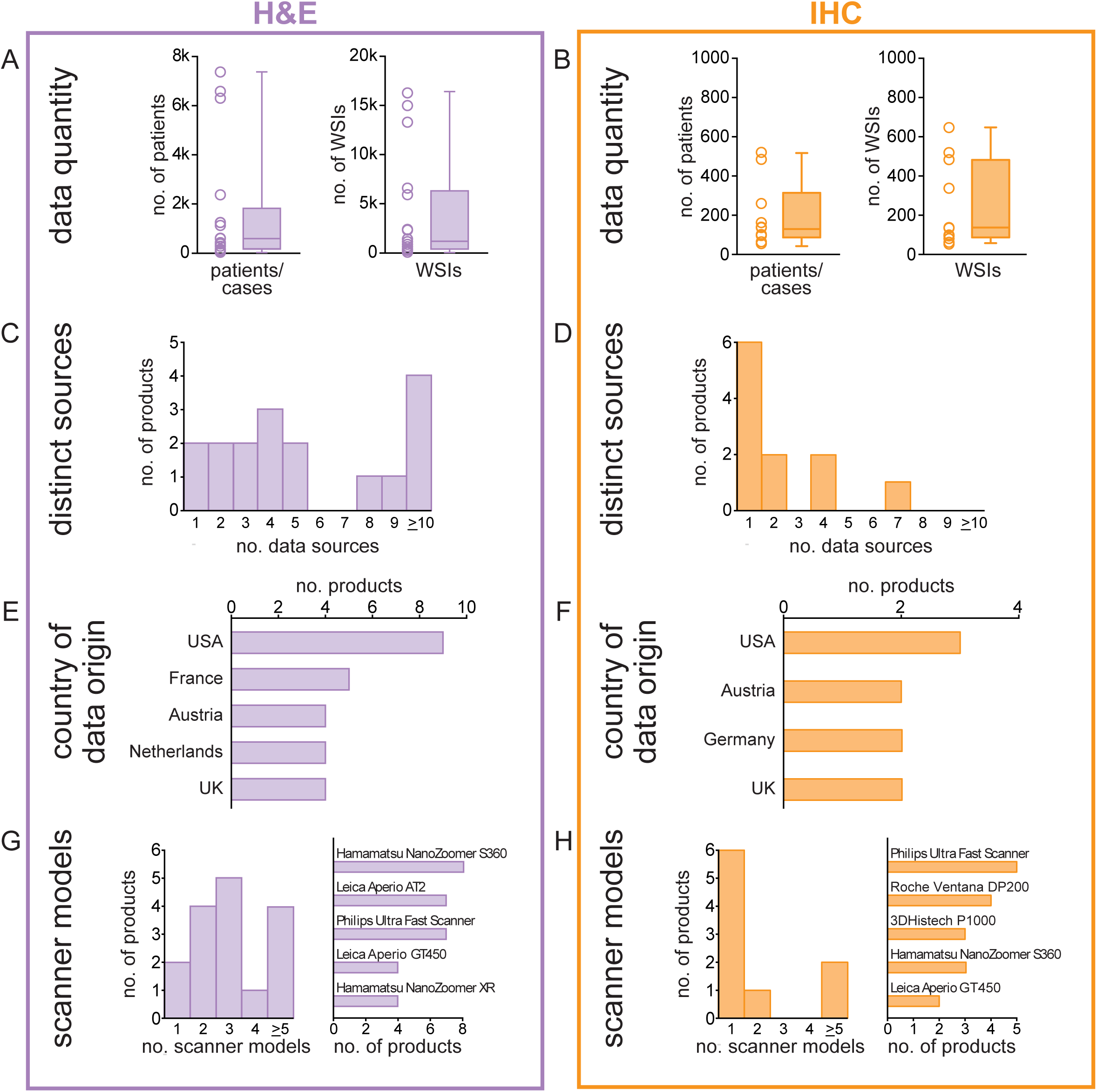
Composition of datasets used in the external validation of AI devices. (A) Box and whisker plots showing the overall size of datasets reported in external validation of AI devices using H&E images and (B) IHC images as input. Each point represents an individual product, for which dataset size has been summed where there were multiple external validation studies available for a single product. Dataset size is reflected by the number of patients/cases used (left panels) and the number of WSIs (right panels), depending on the information available within publications. (C) Histogram indicating the number of distinct data sources included within external validation datasets for each H&E and (D) IHC product (aggregated across multiple studies where appropriate). (E-F) Bar charts showing the countries that were most highly represented in terms of data origin for external validation. (G-H) Histograms (left) showing the number of scanner models used to generate image data for the external validation of each product, and bar charts (right) showing the top five scanner models reported.

Lastly, in terms of dataset composition, we examined the range of scanner models used to generate digital images for external validation datasets, as this is another factor that can introduce variation into digital images and impact AI model performance^27,28^. Consequently, AI models often limit their intended use within regulatory approval to certain scanner models on which they have been validated, even though a range of models are in clinical use across digitised laboratories. When extracting information relating to scanner models used in external validation publications, we found this was reported for 16 H&E products, with 11/16 products reporting external testing overall on 1-3 scanner platforms (Figure 4G). For IHC products, the number of scanner models reported in external validation was lower overall, with 6 of the 9 products that included scanner information only reporting external testing on one scanner model (Figure 4H).

### External validation performance of AI models for cancer detection and grading

Currently, regulatory-approved AI devices to support diagnostic decision-making in digital pathology are not intended to act autonomously and AI outputs require pathologist review before final diagnosis. However, evaluating standalone AI model performance on external datasets (without pathologist oversight) is still an essential step, as this gives an indication of the accuracy of the output that will be displayed to pathologists, which could influence their decision-making. Direct comparisons of the reported performance of AI models in this domain can be challenging for several reasons, even if the models have been designed for a similar intended use. For example, model performance may depend upon the difficulty of the dataset curated for testing, dataset selection criteria, quality control performed on WSIs, ground truth methodology approach^29^, and the performance metrics chosen^30^.

However, in order to provide some insight into the range of performance on external datasets for well-represented AI products (i.e., those for cancer detection/grading), we extracted the reported sensitivity and specificity for models that performed lymph node metastasis, breast cancer, and prostate cancer detection. In addition, the sensitivity, specificity, and quadratic weighted kappa values were extracted from publications on AI models for prostate cancer grading. To understand how these performance metrics compare to pathologists, where possible, we also extracted the reader performance within that study. Overall, reported sensitivity was high (typically >0.9) in most studies, while specificity was more variable but typically >0.8, with reported values generally falling in a similar range to that of pathologists (Figure 5A). Accuracy metrics reported for prostate cancer grading were more variable between studies, but were also broadly similar to pathologists (Figure 5B).

**Figure 5.**
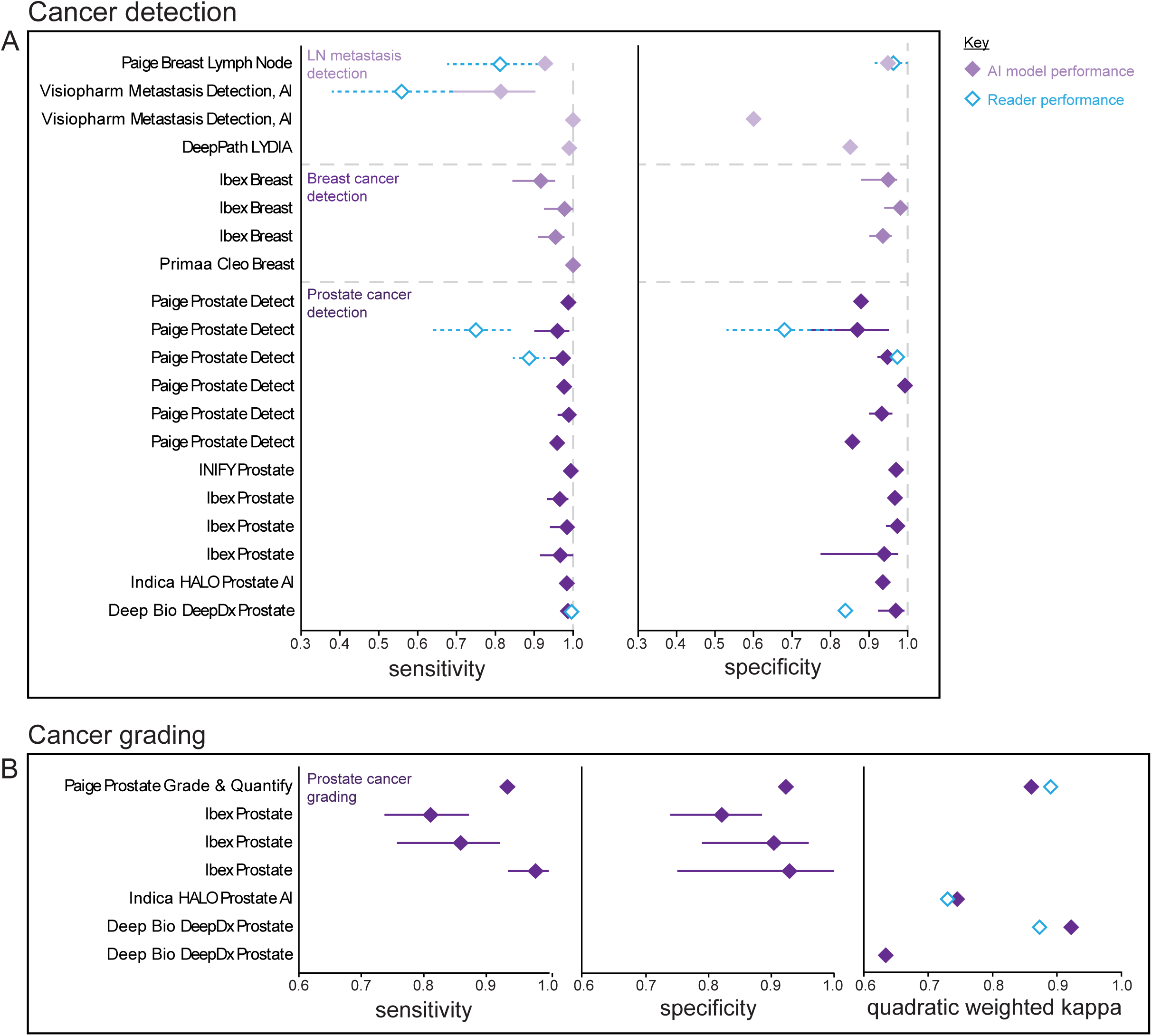
Performance of AI models for cancer detection and grading in external validation studies. (A) Forest plots showing the average standalone AI model sensitivity and specificity reported in each study on each product (with 95% confidence intervals plotted where possible). Products have been grouped into those performing lymph node (LN) metastasis detection, breast cancer detection, and prostate cancer detection. Each line represents a different study on the product indicated. (B) Sensitivity, specificity, and quadratic weighted kappa values reported in external validation studies on AI models performing prostate cancer grading. Values for sensitivity and specificity represent the performance in distinguishing high-grade from low-grade cancer. Open symbols represent the performance of pathologists in studies where this is reported alongside AI model standalone performance.

In external validation studies, AI model performance is typically compared to a reference standard or ground truth, established for each given dataset. However, the approach used to determine ground truth can vary between studies, and group consensus methods are often used for cancer detection/grading given that diagnostic interpretations can differ between pathologists. To appreciate how ground truth has been determined in studies externally validating AI models for cancer detection/grading, we extracted this methodological information from studies where possible (Figure S4A-D). This showed that the most prevalent approach was using the original diagnosis as ground truth, with the remainder of studies reporting using between one and ≥5 readers to determine a consensus ground truth (Figure S4A-D). In 21/29 (76%) of ground truth descriptions, IHC and/or an additional reader was consulted where required. However, when ≥5 readers were used to determine consensus ground truth no additional information was reported as being consulted. These data reflect the diversity in approaches to ground truth determination in studies validating AI model performance, which may be influenced by the information and resources available to the study team.

## Discussion

The mounting enthusiasm that AI-based technologies can deliver improvements to pathology services and patient outcomes needs to be matched by a robust strategy to establish which devices are fit for purpose in real-world clinical workflows. There is already significant clinical resource and infrastructure being dedicated to trialling new healthcare AI products, evaluating their efficacy/effectiveness, and supporting their integration^31–33^. This is, in part, a result of national strategies which have begun to place AI front and centre in the aspirations to transform patient care (e.g.^3,34^). Informed decision-making on AI devices for clinical deployment requires clinicians, patients, and commissioners, to have access to clear, up-to-date, reliable evidence on device performance and clinical utility. Given the substantial time, energy, expertise, resources, and financial input required for local validation, integration, evaluation, and monitoring of AI devices in clinical workflows^35^, the need for robust evidence cannot be overstated.

As medical device regulations and standards evolve, and endeavour to account for the capabilities and risks of AI devices within this rapidly advancing field, it can be difficult to access reliable evidence on existing devices. This can leave clinicians relying on vendor marketing material, and data from non-independent sources, to make purchasing decisions that have the potential to impact patients. Furthermore, the evidence available on regulated devices can vary considerably^36^. In the EU, for example, the extended transition from the old IVDD legislation to the new IVDR, has resulted in a mixture of products on the market that meet the requirements of either the old or new regulations. For AI image analysis products intended to support diagnostic decision-making, the new IVDR assigns a higher risk class than the old IVDD (which allowed manufacturer self-certification), so products now require greater safety/performance evidence, independent review by a Notified Body, and additional post-market monitoring. Therefore, until the transition to IVDR has completed (currently not expected until 2030), disparities will remain in the evidence and requirements for devices on the European market.

### Rigorous validation requires high quality datasets

For AI models, the quality of evidence on their performance is heavily influenced by the quality of data used in their training and testing. In histopathology image analysis, data quality is particular important, due to the range of factors that can introduce variation into digital images, including scanner model/settings^37^, tissue staining protocols^38,39^, dataset size^40^, and population demographics^41^. Robust external validation relies on datasets that reflect the range of factors that may be encountered in clinical workflows, in order to generate performance data that is likely to translate to the real-world^42^. However, our analysis highlights the variability in the diversity incorporated into published validation datasets, which can exacerbate the interpretation of performance metrics reported for AI products. In addition, the use of broad AI accuracy metrics can conceal variability in performance on certain clinical or patient subgroups, if these are poorly represented in the datasets^43^. The chosen metrics may also not always be appropriate to apply to the clinical task in question^30^, or to appropriately convey clinical safety (e.g. applying equivalent weighting to grade differences in prostate cancer diagnosis, when misclassification of certain grades can have a greater impact on patient care than others)^8^. Taken together, this highlights the value of national hidden test datasets for benchmarking AI performance, the need for local validation of AI models, and the necessity to monitor long-term patient outcomes where AI contributes to diagnostic decision-making.

### Product registers and transparency

Despite efforts to improve transparency around medical devices (e.g.,^23,44^), the lack of a centralised source of reliable information on regulatory-approved AI devices has led to several clinical domains launching independent registers in an attempt to bridge this gap (e.g.,^17–21)^. Maintaining an accurate database of product evidence, including vendor-supplied documentation and peer-reviewed publications, can help to counter confusion and inconsistencies that arise with information in the public domain. While conducting our analysis, for example, we noted that the intended use and features of certain devices could differ somewhat, depending on the jurisdiction of regulatory approval. In addition, there were often discrepancies in the product information presented within press releases, regulatory documentation (where available), vendor website descriptions, and third-party platforms, leading to an incomplete understanding of a product’s intended use. Furthermore, when gathering product-related information from online sources, we often could not identify basic information on CE-marked products, e.g., the type of image data or tissue specimen required, or even whether the device utilised an DL/AI-based approach to generate its output.

Another significant challenge we encountered was how to approach changes or updates to existing AI products. For example, if a new version of a product was released or an AI model updated, it was unclear whether the existing clinical evidence in support of the original product could also apply to the updated product. This was further hampered by publications sometimes not clearly stating which version of a product was evaluated (which is a requirement in CONSORT-AI and STARD-AI reporting guidelines^45,46^), and, for some product updates, it was unclear at what point a DL/AI-based approach had been introduced into a device. This highlights an issue that will continue to arise through the deployment and monitoring of AI devices, which will require clear communication of product updates and an understanding of whether existing evidence can legitimately support an updated device. Given that fundamental changes to an AI model can substantially alter performance characteristics, this will be a crucial element of ongoing monitoring.

In spite of the challenges faced in extracting required information, academic peer-reviewed publications remain the most valuable source of information relating to AI model performance. Since conducting our initial analysis of H&E products in 2023, we found that the proportion of products with an associated publication had not increased substantially (remaining at around half of all products), which is a shortcoming noted for AI models in other medical imaging domains^36,47–49^. However, we did locate multiple publications on several CE-marked products, which has allowed us to provide an in-depth analysis using 54 publications from across 29 products, and offer insights into the current standards of evidence quality and product performance in this field.

### Developments within the Pathology AI marketplace since previous analysis in 2023

Our analysis of AI vendors and their products highlighted features of general growth, shifts in focus, and indicators of consolidation. For example, compared with our initial analysis in 2023, we noted instances where vendor websites were no longer available, companies had dissolved or been acquired, and certain products previously listed had been removed. Given that nearly half of the CE-marked devices on the market are manufactured by small companies (with <50 employees), this raises the question of whether products in this domain can be reliably supported and sustained by vendors post-deployment. Concurrently, on the European market, as we edge closer to the end of the IVDR transition period, there may be a reduction in the regulatory-approved products available, if products fail to meet the requirements of the new regulations.

The breadth of vendor offerings, beyond AI devices for approved clinical use, was also noted. Most prominent among these offerings were AI tools directed at biopharmaceutical use rather than clinical diagnostics (e.g., to support drug development through biomarker expression analysis, therapeutic target identification, or optimising patient selection for clinical trials), as well as pan-cancer models designed for broad types of specimen (as opposed to the more narrowly-defined functionality of existing CE-marked devices), and AI based foundation models which can be adapted to numerous types of task. This highlights new challenges in the regulatory space and emphasises the need to monitor market developments.

## Conclusion

Given the vast resource required of clinical teams, hospital IT departments, and technical infrastructure, to integrate, deploy, and monitor AI devices within clinical workflows, robust decision-making on which devices to deploy will require robust evidence. As the number and range of AI products for image analysis in pathology grows, there is an increasing need to generate independent, accessible evidence to show that these products can add value to the clinical workflow, and provide long-term, cost-effective solutions to combat the challenges faced by pathology departments.

## Data Availability

All data analysed in this study was obtained from public sources and is contained within the manuscript.

## Author Contributions

G.A.M. and D.T. designed the study. G.A.M. conducted the searches and G.A.M and L.G. performed data extraction. G.A.M. analysed the data and wrote the manuscript. G.A.M., L.G., C.M., D.B. and D.T. reviewed and edited the manuscript. All authors read and approved the manuscript for publication.

## Acknowledgements

This research is part of the National Pathology Imaging Co-operative, NPIC (Project no. 104687) which was initially supported by a £50 million investment from the Data to Early Diagnosis and Precision Medicine challenge, managed and delivered by UK Research and Innovation (UKRI). We thank Caroline Glover for supporting initial searches and product identification.

## Supplementary Appendix

**Table S1.**
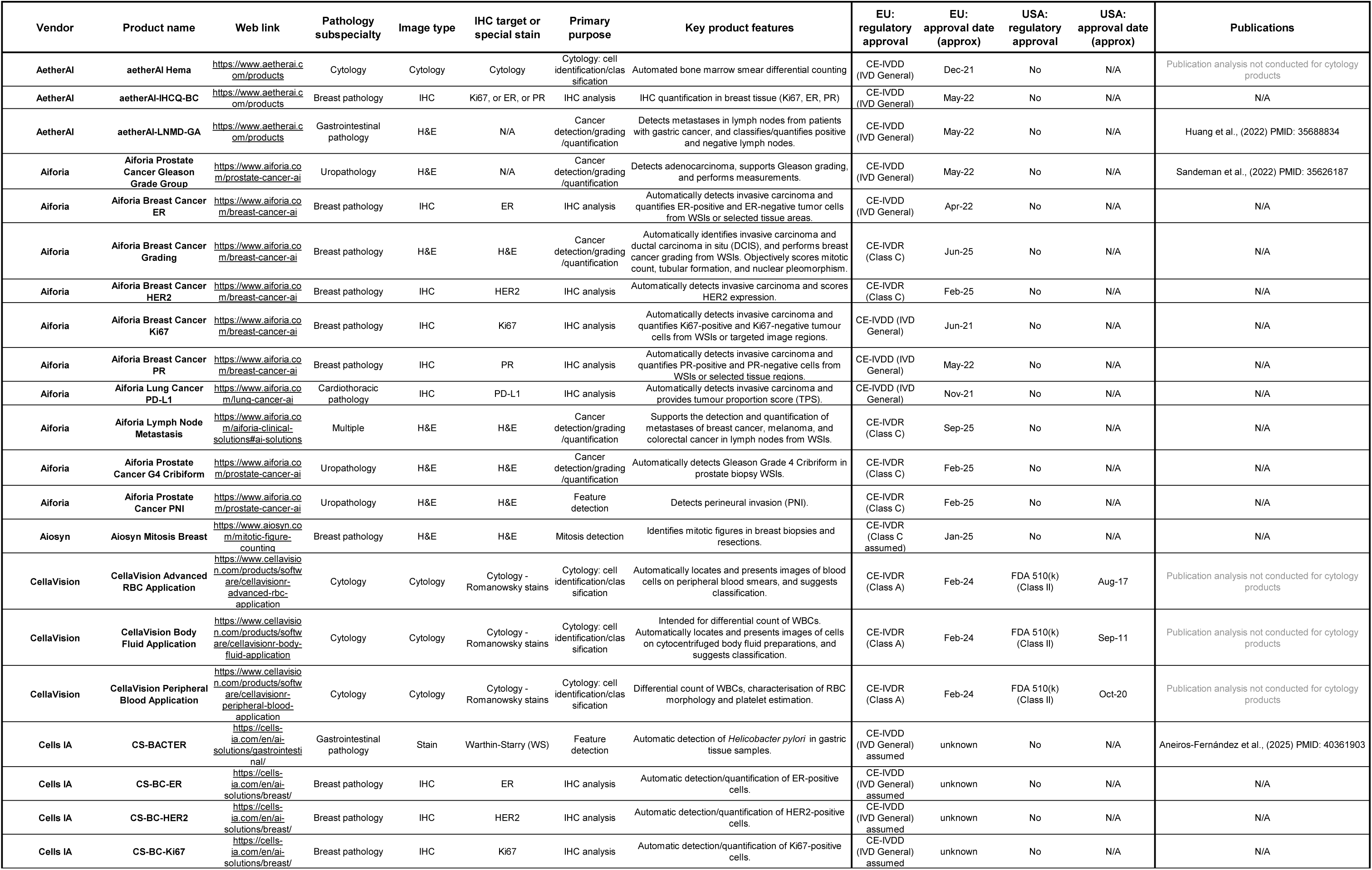
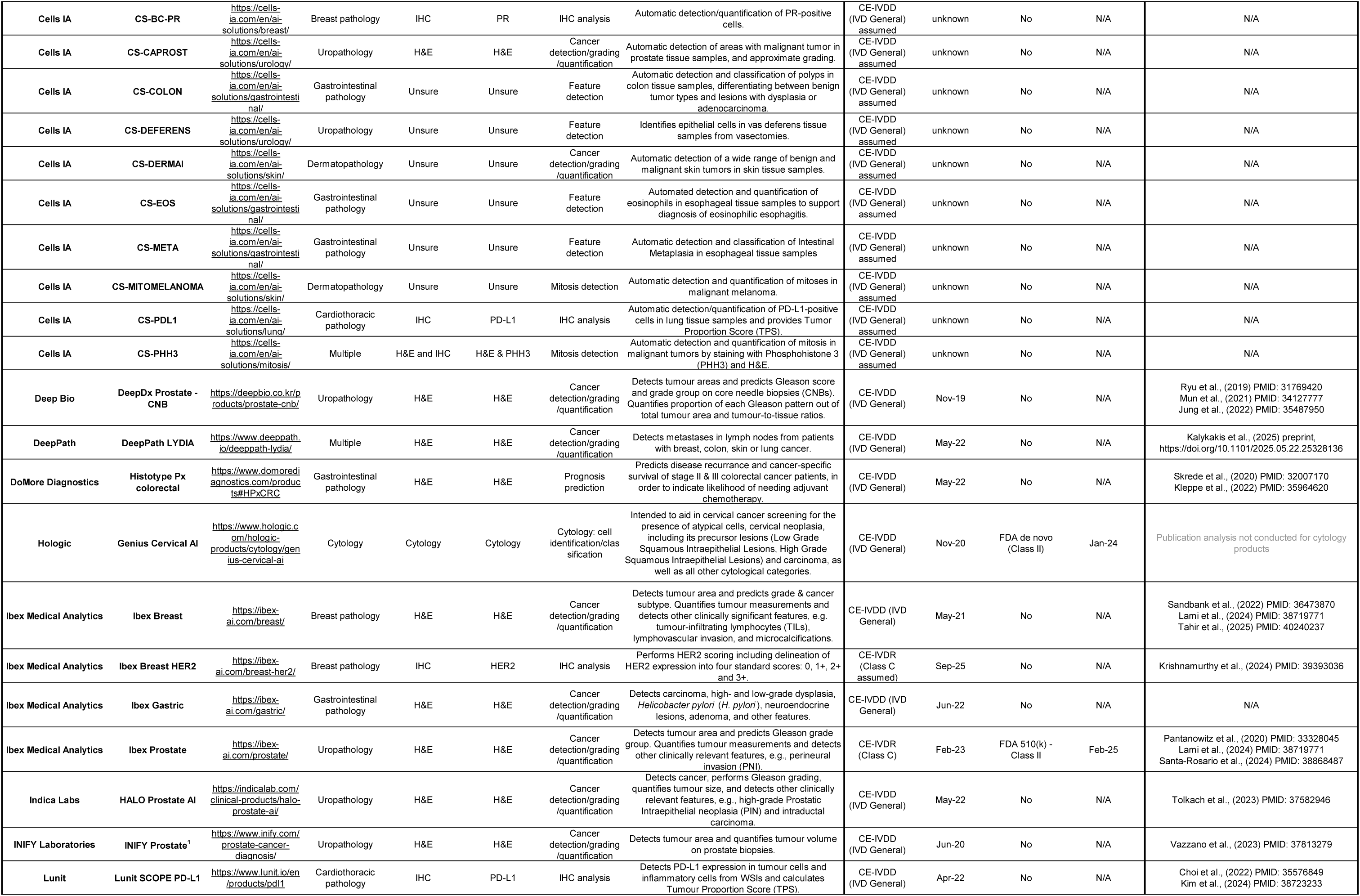
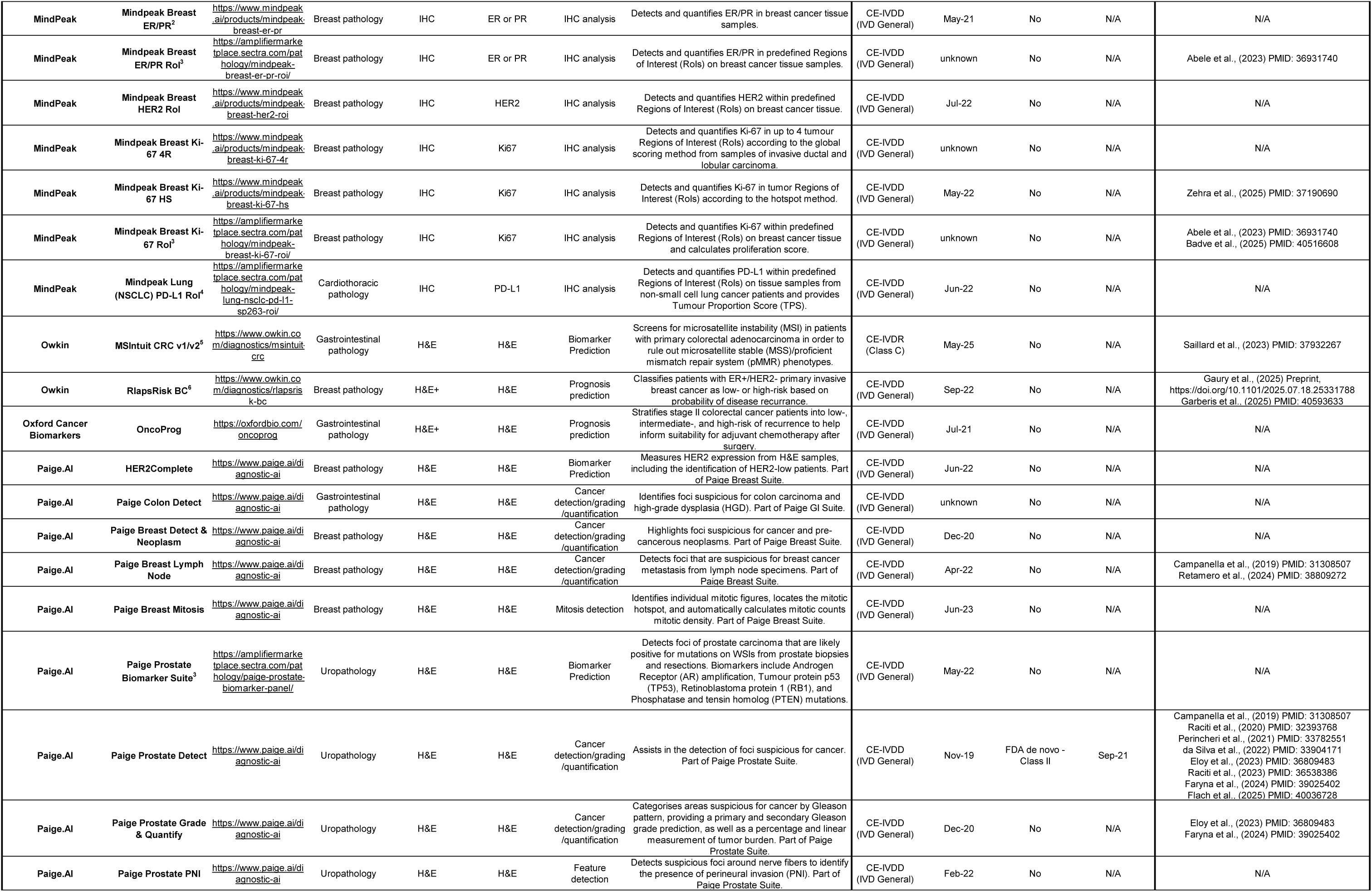
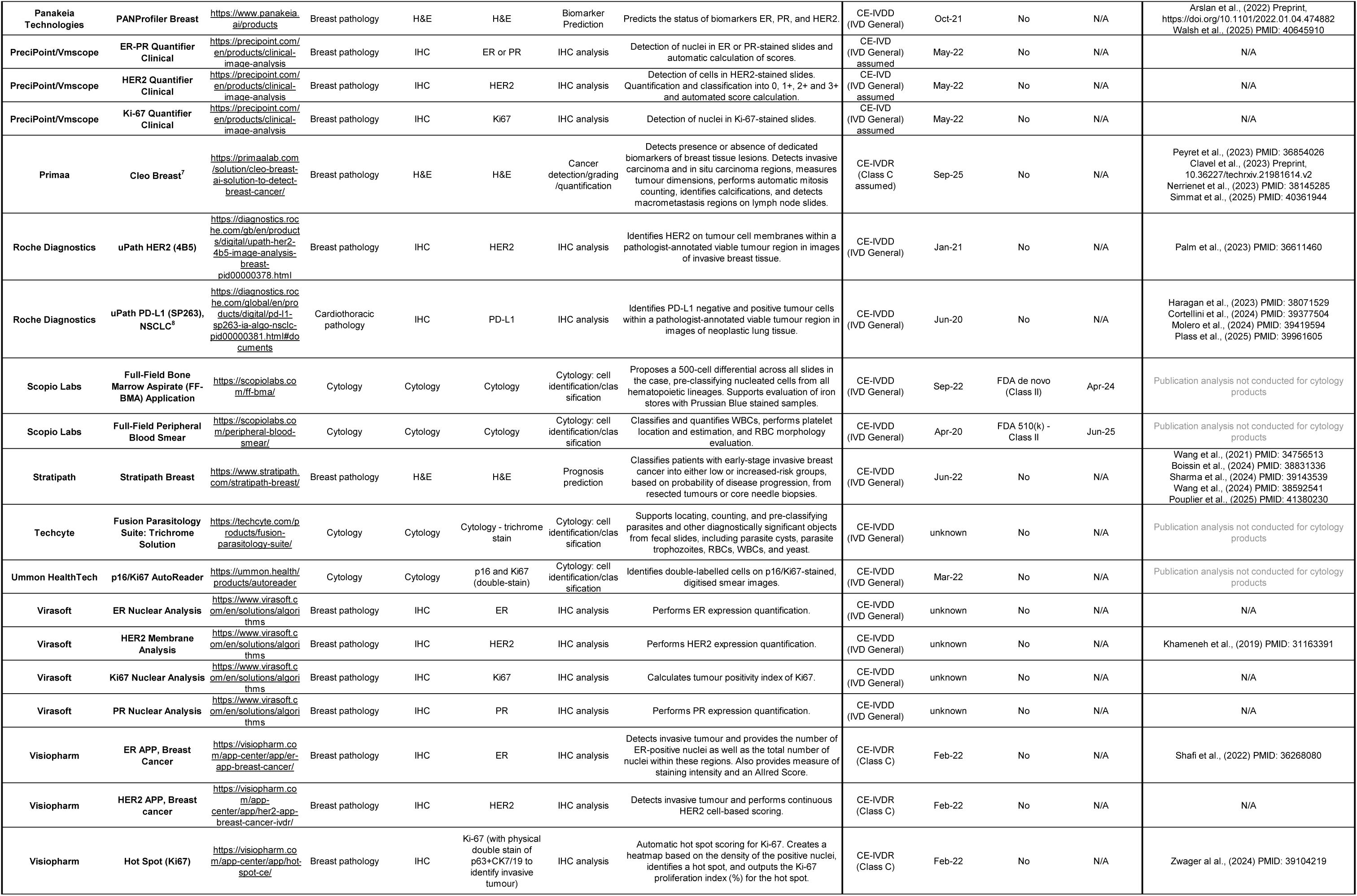
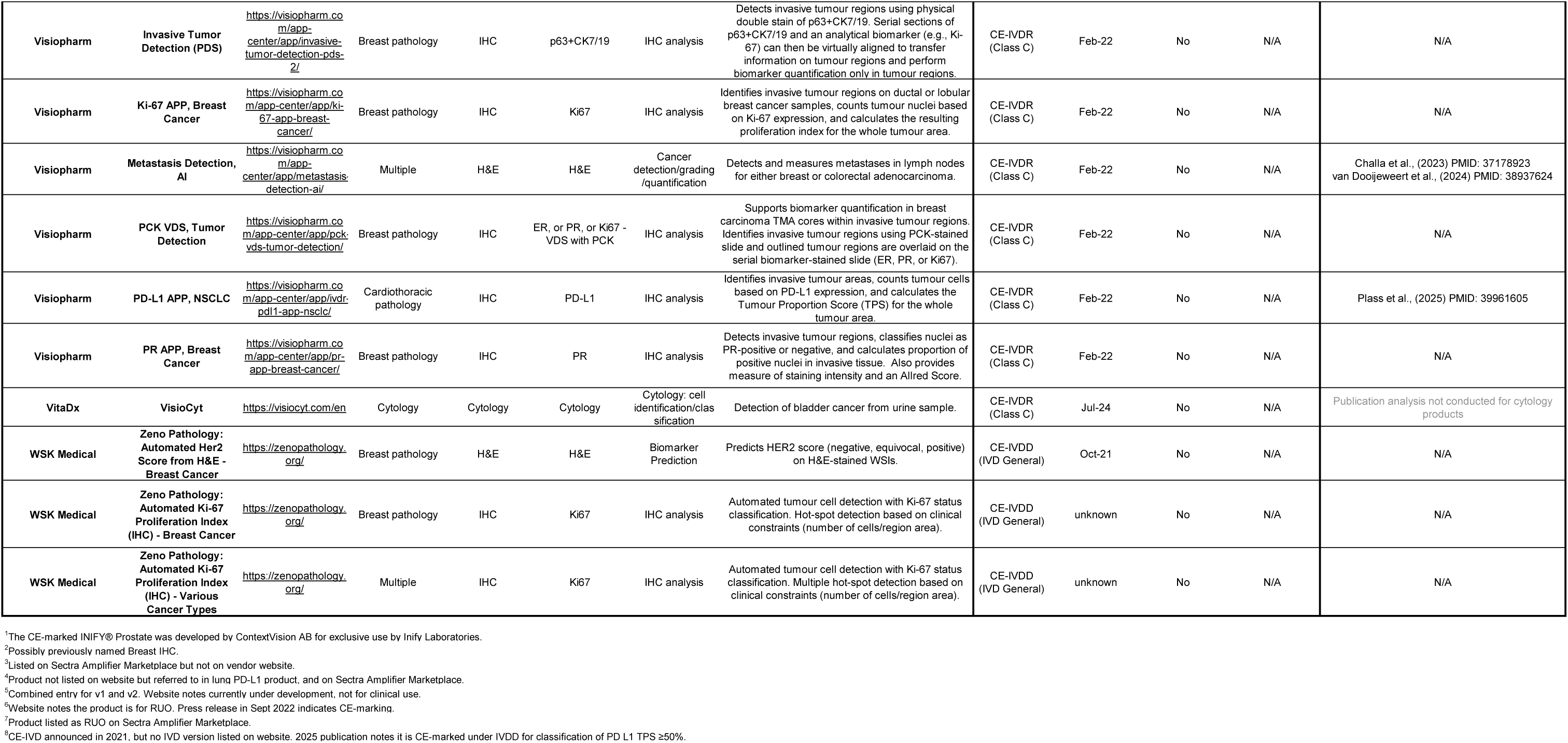
AI devices for image analysis in digital pathology: key features, regulatory approval, and associated publications.

**Figure S1:**
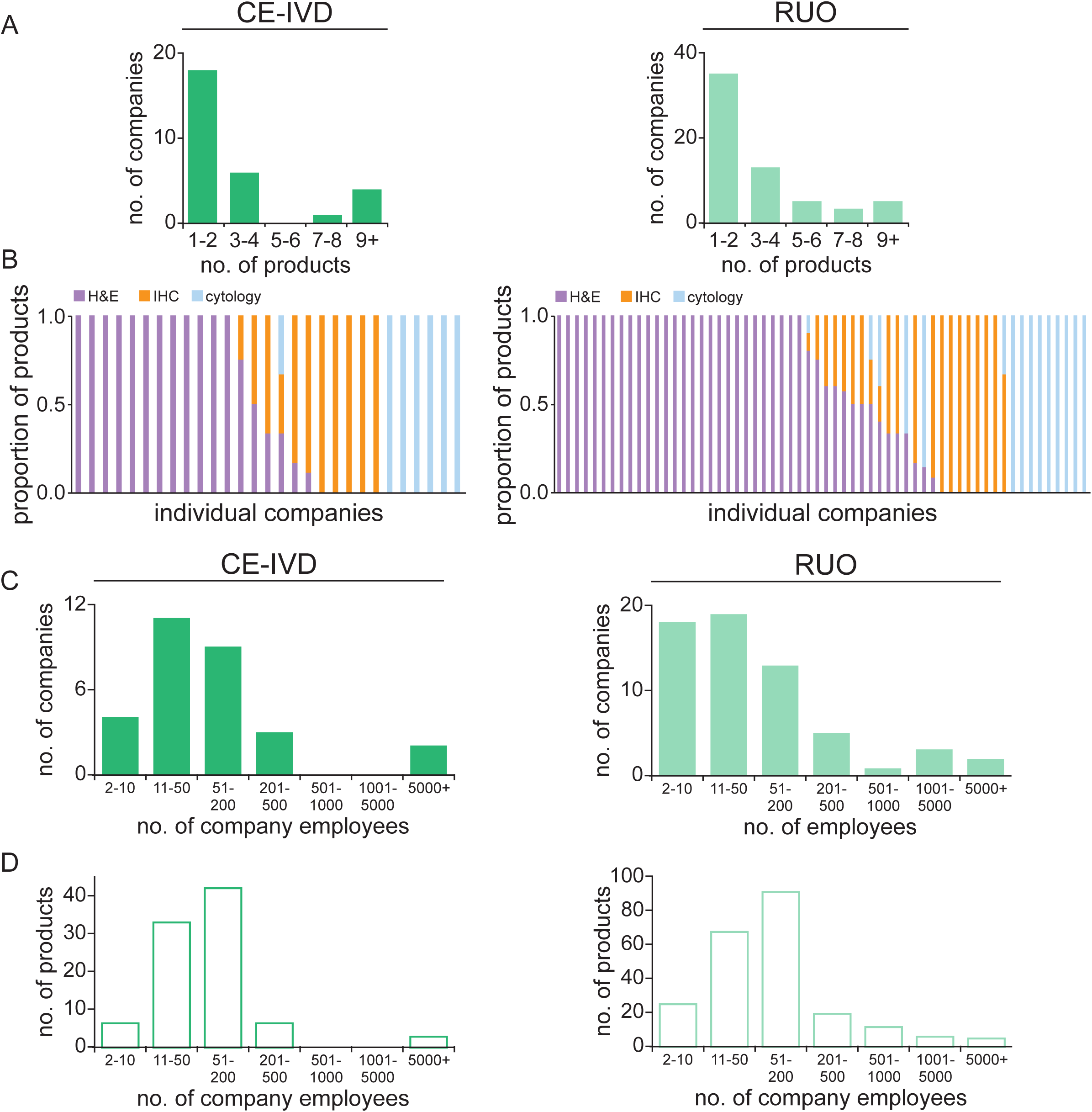
Features of vendors developing AI devices. (A) Bar graphs indicating the number of products developed by each vendor, for CE-marked (left) and RUO products (right), and (B) the image type used as input data for each company’s product(s). (C) Graphs indicating vendor size (as defined by employee number), and (D) the number of products manufactured by vendors in each size categoriy.

**Figure S2:**
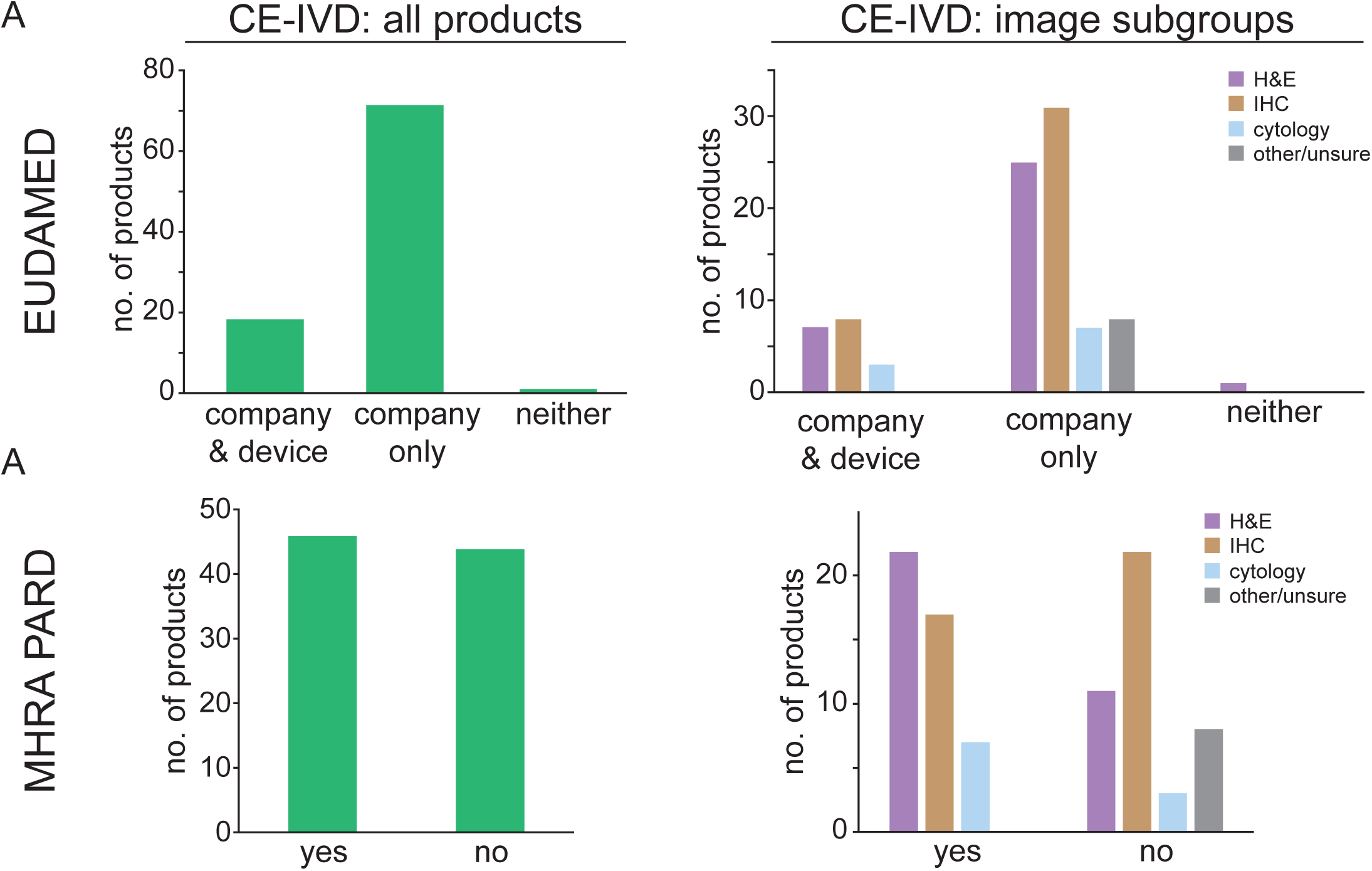
Medical device database listings. (A) Proportion of companies and devices listed on EUDAMED, for all devices reported as having a CE-mark (left) and the distribution by image type (right). (B) Proportion of companies listed on the MHRA PARD, for all devices reported as having a CE-mark (left) and the distribution by image type (right).

**Figure S3:**
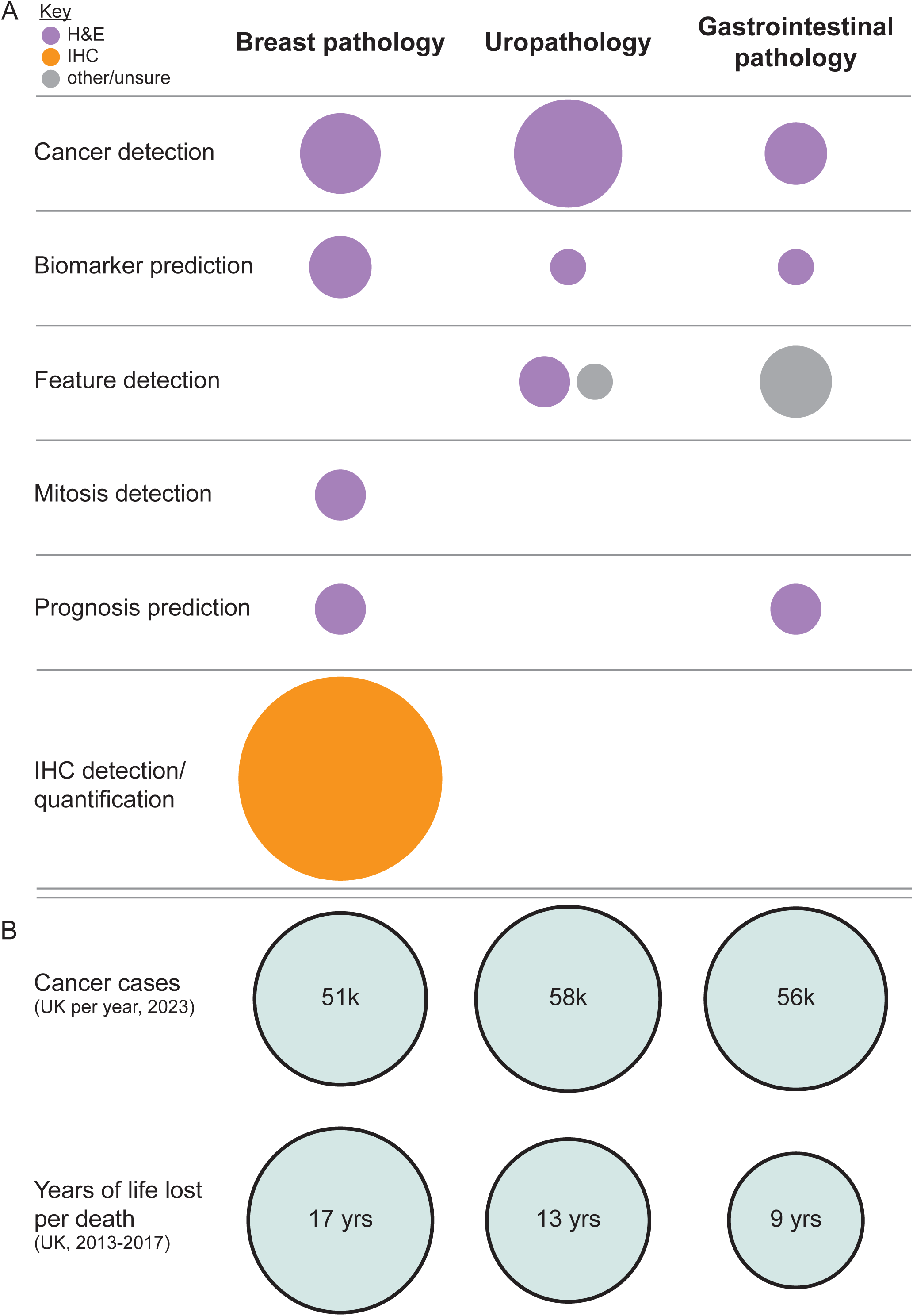
AI devices for image analysis in three clinical subspecialties. (A) Proportional area charts showing the number of CE-marked devices available for each primary function within each clinical specialty, and the image type used as input. (B) Propor-tional area charts showing the number of cancer cases in each subspecialty and the years of life lost per cancer death (based on data from the UK).

**Figure S4:**
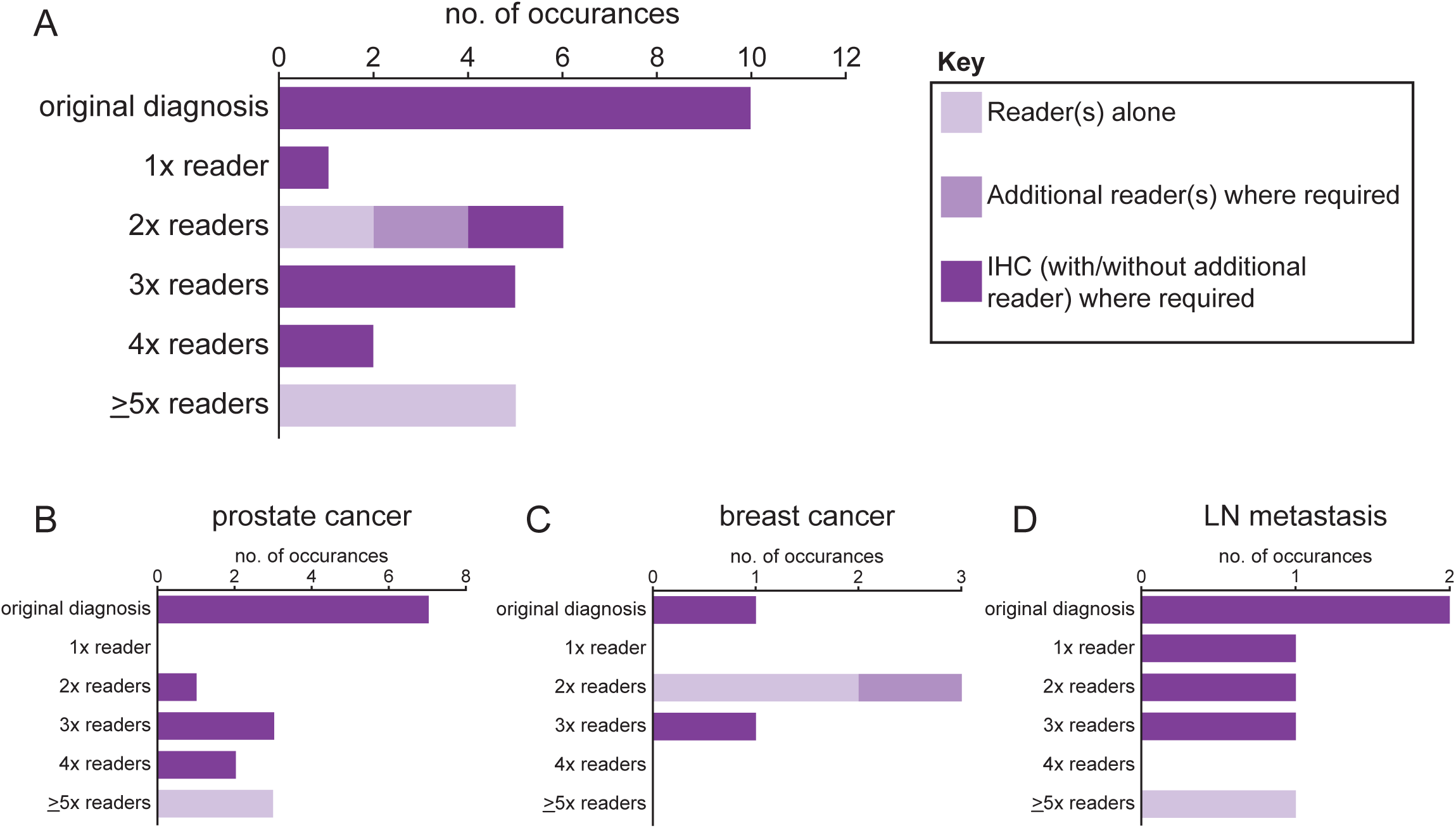
Ground truth methodology approach in external validation studies of AI models for cancer detection/grading. (A) Bar graph showing the number of instances where different approaches to determining the diagnostic ground truth was employed in external validation studies for cancer detection/grading AI models. This includes where ground truth was determined by the original report, or by between 1 and ≥5 readers, with or without additional readers and/or IHC information where required. (B) Approach to ground truth determination used in external validation studies of AI models for prostate cancer, (C) breast cancer, and (D) lymph node metastasis detection.

## References

1. Baxi, V., Edwards, R., Montalto, M. & Saha, S. Digital pathology and artificial intelligence in translational medicine and clinical practice. Mod. Pathol. 35, 23–32 (2022).

2. Rakha, E. A. et al. Current and future applications of artificial intelligence in pathology: a clinical perspective. J. Clin. Pathol. 74, 409–414 (2021).

3. 10 Year Health Plan for England: fit for the future. GOV.UK https://www.gov.uk/government/publications/10-year-health-plan-for-england-fit-for-the-future (2025).

4. Faster, fairer access to HealthTech under new national programme. NICE website: The National Institute for Health and Care Excellence https://www.nice.org.uk/news/articles/faster-fairer-access-to-healthtech-under-new-national-programme (2026).

5. Aristidou, A., Jena, R. & Topol, E. J. Bridging the chasm between AI and clinical implementation. The Lancet 399, 620 (2022).

6. Device Classification Under Section 513(f)(2)(De Novo). https://www.accessdata.fda.gov/scripts/cdrh/cfdocs/cfpmn/denovo.cfm?id=DEN200080.

7. K241232. https://www.accessdata.fda.gov/cdrh_docs/reviews/K241232.pdf.

8. Jia, Y. et al. A deployment safety case for AI-assisted prostate cancer diagnosis. Comput. Biol. Med. 192, 110237 (2025).

9. Cresswell, K. et al. A mixed methods formative evaluation of the United Kingdom National Health Service Artificial Intelligence Lab. NPJ Digit. Med. 8, 448 (2025).

10. Matthews, G. A., McGenity, C., Bansal, D. & Treanor, D. Public evidence on AI products for digital pathology. Npj Digit. Med. 7, 1–11 (2024).

11. McGenity, C. et al. Artificial intelligence in digital pathology: a systematic review and meta-analysis of diagnostic test accuracy. NPJ Digit. Med. 7, 114 (2024).

12. Wagner, S. J. et al. Built to Last? Reproducibility and Reusability of Deep Learning Algorithms in Computational Pathology. Mod. Pathol. 37, 100350 (2024).

13. National Commission into the Regulation of AI in Healthcare. GOV.UK https://www.gov.uk/government/groups/national-commission-into-the-regulation-of-ai-in-healthcare.

14. Regulation (EU) 2024/1689 of the European Parliament and of the Council of 13 June 2024 Laying down Harmonised Rules on Artificial Intelligence and Amending Regulations (EC) No 300/2008, (EU) No 167/2013, (EU) No 168/2013, (EU) 2018/858, (EU) 2018/1139 and (EU) 2019/2144 and Directives 2014/90/EU, (EU) 2016/797 and (EU) 2020/1828 (Artificial Intelligence Act) (Text with EEA Relevance). (2024).

15. U.S. Food and Drug Administration. Artificial Intelligence-Enabled Device Software Functions: Lifecycle Management and Marketing Submission Recommendations. https://www.fda.gov/regulatory-information/search-fda-guidance-documents/artificial-intelligence-enabled-device-software-functions-lifecycle-management-and-marketing (2025).

16. Ong, A. Y. et al. Current challenges and the way forwards for regulatory databases of artificial intelligence as a medical device. Npj Digit. Med. 10.1038/s41746-026-02407-w (2026) doi:10.1038/s41746-026-02407-w.

17. Ambient Voice Technology Self-Certified Supplier Registry. NHS Transformation Directorate https://transform.england.nhs.uk/digitise-connect-transform/digitising-the-frontline/ambient-voice-technology-self-certified-supplier-registry/.

18. AI Registry Listing | The Royal College of Radiologists. https://www.rcr.ac.uk/our-services/artificial-intelligence-ai/ai-registry/.

19. AI directory. The Royal College of Ophthalmologists https://www.rcophth.ac.uk/ai-directory/.

20. Silkens, M. E. W. M., Ross, J., Hall, M., Scarbrough, H. & Rockall, A. The time is now: making the case for a UK registry of deployment of radiology artificial intelligence applications. Clin. Radiol. 78, 107–114 (2023).

21. ACR - AI Central. https://aicentral.acrdsi.org//.

22. Products. Health AI Register https://healthairegister.com/pathology/products.

23. EUDAMED database - EUDAMED. https://ec.europa.eu/tools/eudamed/#/screen/home.

24. PARD. https://pard.mhra.gov.uk/.

25. Cancer Registration Statistics, England, 2023. NHS England Digital https://digital.nhs.uk/data-and-information/publications/statistical/cancer-registration-statistics/england-2023.

26. Ahmad, A. S. et al. Years of life lost due to cancer in the United Kingdom from 1988 to 2017. Br. J. Cancer 129, 1558–1568 (2023).

27. Aubreville, M. et al. Mitosis domain generalization in histopathology images -- The MIDOG challenge. Med. Image Anal. 84, 102699 (2023).

28. Thiringer, E., Gustafsson, F. K., Eriksson, K. L. & Rantalainen, M. Scanner-Induced Domain Shifts Undermine the Robustness of Pathology Foundation Models. Preprint at 10.48550/arXiv.2601.04163 (2026).

29. Foucart, A., Debeir, O. & Decaestecker, C. Shortcomings and areas for improvement in digital pathology image segmentation challenges. Comput. Med. Imaging Graph. 103, 102155 (2023).

30. Maier-Hein, L. et al. Metrics reloaded: recommendations for image analysis validation. Nat. Methods 21, 195–212 (2024).

31. World-leading AI trial to tackle breast cancer launched. GOV.UK https://www.gov.uk/government/news/world-leading-ai-trial-to-tackle-breast-cancer-launched (2025).

32. NHS England. NHS Accelerated Access Collaborative » Artificial Intelligence in Health and Care Award. https://www.england.nhs.uk/aac/what-we-do/how-can-the-aac-help-me/ai-award/.

33. Bold bet on AI to keep UK at forefront of science and research breakthroughs from healthcare, to better public services. GOV.UK https://www.gov.uk/government/news/bold-bet-on-ai-to-keep-uk-at-forefront-of-science-and-research-breakthroughs-from-healthcare-to-better-public-services.

34. Baumgart, D. C. & Kvedar, J. C. Germany and Europe lead digital innovation and AI with collaborative health data use at continental level. Npj Digit. Med. 8, 215 (2025).

35. Wilkinson, L. S., Dunbar, J. K. & Lip, G. Clinical Integration of Artificial Intelligence for Breast Imaging. Radiol. Clin. North Am. 62, 703–716 (2024).

36. Chouffani El Fassi, S., et al. Not all AI health tools with regulatory authorization are clinically validated. Nat. Med. 30, 2718–2720 (2024).

37. Humphries, M. P. et al. Development of a multi-scanner facility for data acquisition for digital pathology artificial intelligence. J. Pathol. 264, 80–89 (2024).

38. Swiderska-Chadaj, Z. et al. Impact of rescanning and normalization on convolutional neural network performance in multi-center, whole-slide classification of prostate cancer. Sci. Rep. 10, 14398 (2020).

39. Dunn, C., Brettle, D., Hodgson, C., Hughes, R. & Treanor, D. An international study of stain variability in histopathology using qualitative and quantitative analysis. J. Pathol. Inform. 17, 100423 (2025).

40. Riley, R. D. et al. Importance of sample size on the quality and utility of AI-based prediction models for healthcare. *Lancet Digit*. Health 7, 100857 (2025).

41. Homeyer, A. et al. Recommendations on compiling test datasets for evaluating artificial intelligence solutions in pathology. Mod. Pathol. 35, 1759–1769 (2022).

42. Liu, X. et al. The medical algorithmic audit. *Lancet Digit*. Health 4, e384–e397 (2022).

43. Evans, H. & Snead, D. Understanding the errors made by artificial intelligence algorithms in histopathology in terms of patient impact. NPJ Digit. Med. 7, 89 (2024).

44. U.S. Food and Drug Administration. Artificial Intelligence-Enabled Medical Devices. FDA https://www.fda.gov/medical-devices/software-medical-device-samd/artificial-intelligence-enabled-medical-devices (2026).

45. Sounderajah, V. et al. The STARD-AI reporting guideline for diagnostic accuracy studies using artificial intelligence. Nat. Med. 31, 3283–3289 (2025).

46. Liu, X., Cruz Rivera, S., Moher, D., Calvert, M. J. & Denniston, A. K. Reporting guidelines for clinical trial reports for interventions involving artificial intelligence: the CONSORT-AI extension. Nat. Med. 26, 1364–1374 (2020).

47. Ebrahimian, S. et al. FDA-regulated AI Algorithms: Trends, Strengths, and Gaps of Validation Studies. Acad. Radiol. 29, 559–566 (2022).

48. van Leeuwen, K. G., Schalekamp, S., Rutten, M. J. C. M., van Ginneken, B. & de Rooij, M. Artificial intelligence in radiology: 100 commercially available products and their scientific evidence. Eur. Radiol. 31, 3797–3804 (2021).

49. Antonissen, N. et al. Artificial intelligence in radiology: 173 commercially available products and their scientific evidence. Eur. Radiol. 36, 526–536 (2026).

